# Differences in COVID-19 cyclicity and predictability among U.S. counties and states reflect the effectiveness of protective measures

**DOI:** 10.1101/2022.09.28.22280465

**Authors:** Claudio Bozzuto, Anthony R. Ives

## Abstract

Data available for COVID-19 in the USA make it possible to assess the dynamics of disease spread with 20:20 hindsight. Here, we analyze archived data to explain variation among counties and states in the cyclicity and predictability (that is, the extent to which predictions are possible) of disease dynamics, using a combination of statistical and simulation models. For the period after the initial outbreak but before widespread vaccination (May 2020 – February 2021), we show that for half the counties and states the spread rate of COVID-19, *r*(*t*), was predictable at most 9 weeks and 8 weeks ahead, respectively, corresponding to at most 40% and 35% of an average cycle length of 23 weeks and 26 weeks. However, there were large differences among counties and states, and high predictability was associated with high cyclicity of *r*(*t*). Furthermore, predictability was negatively associated with *R*_0_ values from the pandemic’s onset. This suggests that a severe initial outbreak induced strong and sustained protective measures to lower disease transmission, and these protective measures in turn reduced both cyclicity and predictability. Thus, decreased predictability of disease spread should be viewed as a by-product of positive and sustained steps that people take to protect themselves and others.

**Significance statement:** During the COVID-19 pandemic, many quantitative approaches were employed to predict the course of disease spread. However, forecasting faces the challenge of inherently unpredictable spread dynamics, setting a limit to the accuracy of all models. For counties and states in the USA, we document very high variation in predictability after the initial outbreak and before widespread vaccination. Jurisdictions with high predictability were those that showed pronounced cyclic re-emergences (‘waves’). The variation in predictability can be explained by differences in the human responses to disease: jurisdictions in which individuals and authorities took strong and sustained protective measures against COVID-19 successfully curbed subsequent waves of disease spread, but at the same time unintentionally decreased its predictability.

## Introduction

Human societies have always experienced outbreaks of infectious diseases, and disease epidemics are expected to emerge or re-emerge more frequently in the future (1–3). The COVID-19 pandemic, caused by the SARS-CoV-2 virus, showed the limited strategies and actions humans have at their disposal to prevent outbreaks of emerging diseases, and the suffering and death once a disease starts spreading (2, 4, 5).

If a disease outbreak cannot be prevented, public health officials and politicians will try to swiftly implement measures to help minimize disease-related suffering and death (6, 7). Such measures can range from preparing and re-organizing medical infrastructure (e.g., increasing personnel for intensive care units) to enacting non-pharmaceutical interventions (NPIs), either as mandates or as recommendations to the public. For impending or unfolding disease outbreaks, forecasts have proven helpful for emergency planning (7, 8). To match the time required to plan and implement mitigation actions for public health needs, however, the lead-time of the forecasts typically ranges from one week to two or more months (7, 9). Long-term forecasts are important to prepare for resurgences of the disease, as has happened worldwide with COVID-19 (10, 11), and also to justify severe NPI mandates such as lockdowns: mandates that disrupt social and economic systems can be justified if the course of the disease spread is expected to last months and lead to a high death toll. For re-emerging influenza outbreaks, Viboud and Vespignani (9, p. 2804) aptly use a weather forecast analogy: “the influenza forecasting community will need to offer weather forecasts as well as climate predictions.”

The COVID-19 pandemic has spurred an unprecedented effort to quantitatively understand disease spread and forecast spread dynamics to help public health officials implement protective measures such as NPIs (12, and references therein). Nonetheless, these efforts face the challenge that the predictability of COVID-19 spread may be inherently limited. Here, we use the definition that “predictability is the study of the extent to which events can be predicted” (13, p. 2425). Several epidemiological studies have addressed the fundamental limit to predictability of disease spread using model-free, entropy-based approaches (14–16). For example, Scarpino and Petri (15) found that for nine human diseases, there is a barrier to predictability, but that single outbreaks are in general predictable and that predictability depends in part on the basic reproduction number, *R*_0_. Furthermore, these authors found considerable variation in predictability among jurisdictions for single diseases. In comparison, assessments of realized predictability, i.e. forecast accuracy, for influenza and COVID-19 outbreaks have shown that four weeks seems to be the forecast horizon beyond which the dynamics are hard to predict (9, 17–19), implying that predicting COVID-19 resurgences two months in advance may be futile.

Model-free approaches address predictability with methods heavily relying on information theory. We worry that public health officials facing an epidemic and planning for public health responses need more concrete assessment of the limits to predictability as well as the factors that might determine this predictability. Here, we use time series models to statistically fit disease spread dynamics, and then analyze the predictability of the fitted models using the measure predictive power, *PP*(*t*), rooted in information theory and developed in climatology (20) (see also (13)). An advantage of our approach is that we can associate predictability to specific dynamical patterns observed during the pandemic, like cyclic dynamics, which potentially lead to more accurate predictions (e.g. 21, 22).

For centuries it has been known that infectious disease outbreaks resurge regularly over time (see e.g. (23) for historical background). Resurgent outbreaks can have many causes such as seasonality, school terms, or new pathogen variants (23, 24, 25, and references therein). For COVID-19, too, the dynamics are characterized by ‘waves’ or cycles, not only in the USA but throughout the world, and different cyclic patterns have been documented, for example, at weekly and seasonal time scales (10, 11, 22). Moreover, for many countries in both hemispheres additional cycles occur with a period of approximately 4 months (3–6 months), similar to other communicable (viral) diseases like the Spanish flu from 1918 (approximately 5 months; (11)). Mitchell and Zhang (11) speculate that these cycles are caused by virus-host feedbacks, and other studies show that models incorporating behavioral responses to limit disease spread can show cyclic dynamics when these responses occur with a time delay (26–28). We investigate the cyclic dynamics of COVID-19 using a stochastic epidemiological model to understand how human responses to infection rates may affect cyclicity and predictability of disease spread. Specifically, we ask what factors might underlie differences in cyclic dynamics among counties and states in the USA.

Our overall goal is to understand the high variation among counties and states in predictability of COVID-19 spread dynamics during the period after its establishment (May 2020) and before vaccinations became widely available (February 2021). We use this variation to develop an explanation for cyclicity and predictability of the COVID-19 pandemic.

## Methods

### Estimation of COVID-19 spread rate r(t)

We base our analyses on the disease spread rate, *r*(*t*), of COVID-19 in the USA, estimated at the county and state levels (henceforth jurisdictions) using weekly death counts (29) from 9 May 2020 to 12 February 2021 (40 weeks). We did not consider the initial outbreak (March-early May 2020) because there was pronounced among-jurisdiction variation in the time of onset (30), and because protective measures (individual behavior and NPIs) built up differently during the first outbreak (31). We ended the data on 12 February 2021 because vaccinations had started to influence the disease transmission and death rates (32). For *r*(*t*), we used the weekly difference between two adjacent log-transformed death counts; thus, at the original scale death count *D*(*t*) ∝*D* (*t* −1) exp (*r*(*t* − 1)). We used death counts rather than reported cases of disease because death data are less likely to give biased estimates of spread rates than case data (33). Furthermore, predicting death rates is critical for health care in terms of both direct human costs and medical preparedness for increases in critical cases of infection. At the state level, we used data for the 49 conterminous states in the USA (including the District of Columbia), while at the county level we selected the 100 counties in these states with the highest population size to maximize estimation accuracy. To estimate *r*(*t*) using the entire time series, we used a time-varying autoregressive model (30) with (i) death counts as observations, (ii) two process equations capturing the dynamics of the ‘true’ death count and the latent variable *r*(*t*), respectively, and (iii) state-dependent error terms. After fitting the model using the Kalman filter, we used the Kalman smoother (34) to generate jurisdiction-specific time series of *r*(*t*). Figure 1 shows example data and estimated *r*(*t*) time series of three counties, and the *SI Appendix* (Fig. S1-S2) shows all estimated *r*(*t*) time series at the county and state levels grouped by similarity of the spread dynamics. These fits of *r*(*t*) are the best 20:20 hindsight estimates that use all data in the time series. For real-time forecasting, short time series will cause uncertainty in model parameter estimates, but because we are interested in the inherent limit to predictability of the process underlying *r*(*t*), we use the best estimates possible from the entire time series.

**Fig. 1.**
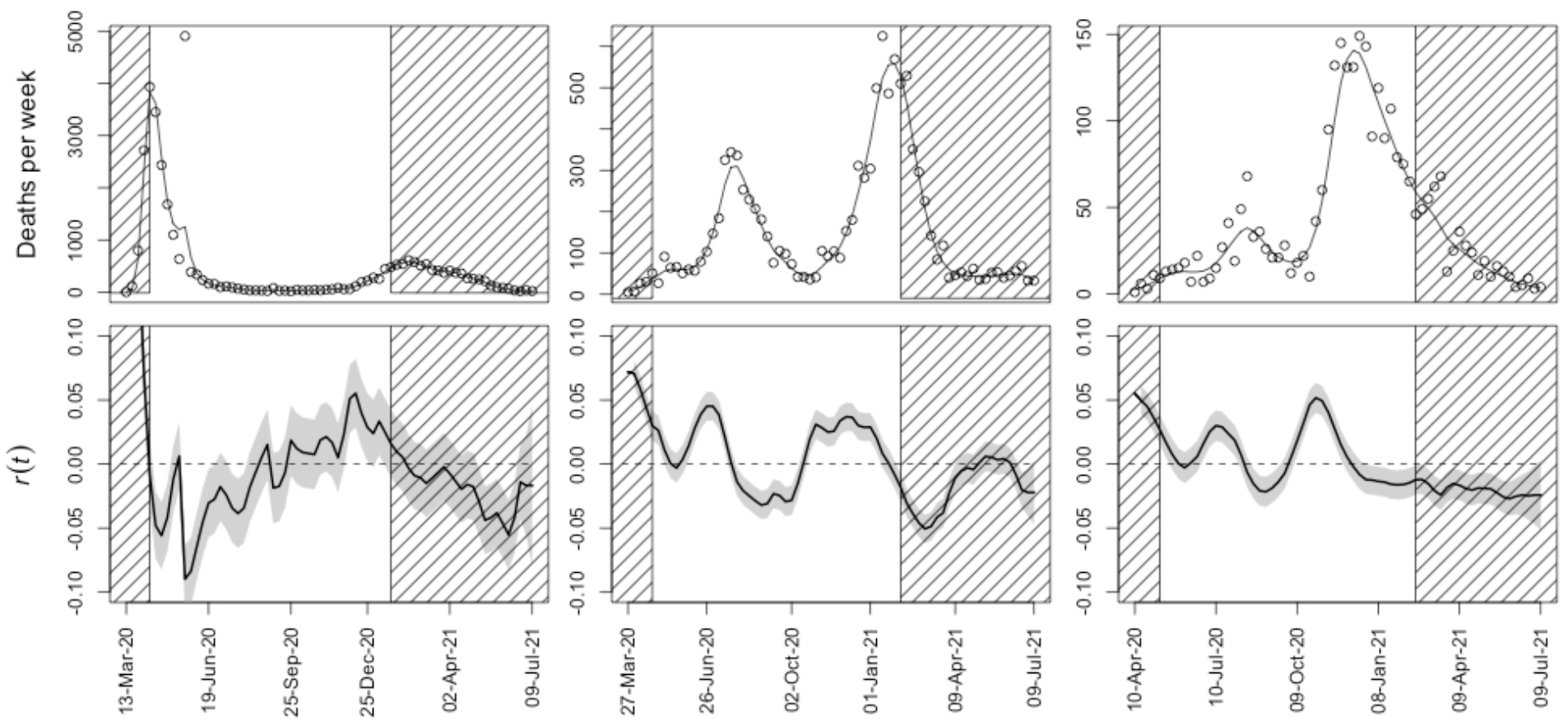
Death data from three illustrative counties in the USA and estimated disease spread rates. Weekly death count data and resulting estimates of *r*(*t*) are given for **(a**,**d)** New York (five boroughs), New York, **(b**,**e)** Maricopa County, Arizona, and **(c**,**f)** El Paso County, Texas. In panels (a-c), points give the data from (29), and the lines give the fit from the Kalman smoother. Panels (d-f) give the corresponding estimates of *r*(*t*) from the Kalman smoother, with the shaded band encompassing the 66% confidence interval. Values of *r*(*t*) within the hatched region at the start and end of the time series were removed for our analyses to exclude the initial outbreak and possible effects of vaccination on the dynamics.

### Analysis of estimated r(t) time series

To analyze the estimated county- and state-level *r*(*t*) time series, we used an autoregressive moving-average (ARMA) time-series model (35, 36). Because *r*(*t*) depends on the difference in death counts between weeks, analyzing *r*(*t*) is qualitatively similar to analyzing death counts as a autoregressive integrated moving average ARIMA(*p*,1,*q*) process of log-transformed deaths counts (see e.g. (10)). We fit a spatial ARMA(2,2) model to both county- and state-level datasets separately, in which each jurisdiction had its own autoregressive coefficients, but all jurisdictions shared the same moving average coefficients, and random errors were assumed to be spatially autocorrelated. We chose the AR order *p* = 2 because it is a parsimonious choice to produce and fit cyclic dynamics (37). We chose the MA order *q = p* = 2 to implicitly account for observation error (38). The model for every jurisdiction *i* is

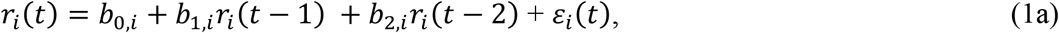

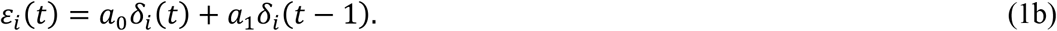

Here, *r*_*i*_(*t*)is the spread rate in jurisdiction *i* for week *t, b*_0,*i*_ gives differences in the mean spread rate among jurisdictions, *b*_1,*i*_ and *b*_2,*i*_ give the jurisdiction-specific AR coefficients for lag-1 and lag-2, *a*_0_ and *a*_1_ are the MA coefficients for lag-0 and lag-1, and *δ*_*i*_ (*t*)is a multivariate Gaussian random variable that incorporates spatial correlation. Spatial correlation between two jurisdictions *i* and *j* is given by cor (*δ*_*i*_(*t*), *δ*_*j*_(*t*))=(1 − *η*)exp (−(*d*_*i,j*_*ϱ*^−1^)^2^), where *d*_*i,j*_is the distance between the two jurisdictions, *η* is the nugget, and *ϱ* is the range (39); parameters *η* and *ϱ* were estimated along with the AR and MA coefficients.

### Cyclic dynamics

The potential cyclicity of the dynamics given by eq. 1 depends on the estimated ARMA model parameters *b*_1_ and *b*_2_ (e.g. 37). For a stationary oscillatory process, the average cycle length (henceforth period) is 2*πw*^−1^, where tan (*w*)=(|*b*_1_^2^ +4*b* |)^1/2^*b*_1_^−1^. We further use the damping factor *d* to characterize cyclicity; *d*scales with the rate at which the amplitude of the cycle decreases over time in the absence of stochasticity. This can be seen in the explicit solution *r*(*t*)=*d*^*t*−1^ (*r*_1_sin (*tw*)− *dr*_0_sin ((*t* − 1)*w*))sin^−1^ (*w*), where *r*_0_ and *r*_1_ are the initial values of *r t* at time point 0 and 1, respectively, and *d*is the damping factor; for a stationary process, *d*<1, and values close to zero imply rapid decreases in amplitude. The damping factor can be expressed in terms of the autoregressive lag-2 coefficient as *d*^2^ =−*b*_2_.

### Predictive power

To assess predictability, we use the measure predictive power, *PP*(*t*)(20), defined as the time-dependent variance of the transition distribution scaled by the variance of the stationary distribution of the ARMA(2,2) process (*SI Appendix*, Fig. S3). If both variances are equal, then no information is available for a forecast to be ‘better’ than a randomly drawn process state according to the stationary distribution, and therefore predictability is said to be lost (13). Because the transition and stationary distributions are properties of the underlying processes that generate stochastic dynamics, *PP*(*t*)gives the theoretical limit of the predictive ability of any model fit to the data.

For a general multivariate Gaussian process, *PP* (*t*)is defined as

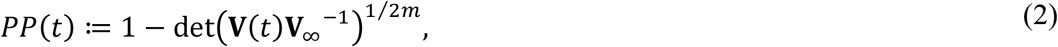

where det(·)*i*s the determinant, **V**(*t*)and **V**_∞_ are the covariance matrices of the transition and stationary distributions, and *m* is the dimension; calculation of **V** (*t*)and **V**_∞_ is outlined in the *SI Appendix*. Because our ARMA(2,2) model (eq. 1) is a univariate process, **V** (*t*)and **V**_∞_ are scalars and *m*=1. Here, *PP*(*t*)can be related to the theoretical limit of forecast accuracy: if *R*^2^ *τ* denotes the coefficient of determination of a predicted value of *r*(*τ*)(*τ* weeks into the future), then the maximum possible value of *R*^2^ (*τ*)is 1 −(**V** (*τ*)**V**_∞_ ^−1^)=1 − (1 −*PP* (*τ*))^2^.

The time dependency of *PP*(*t*)implies a monotonic decrease in predictability with time, eventually approaching zero. Although the approach to zero is usually defined as the predictability barrier (13), from an empirical perspective we set the threshold using the link between prediction *R*^2^ and *PP*(*t*)as follows. As a rule of thumb, values of prediction *R*^2^ <0.25 can be considered as reflecting a very weak match between true and forecasted dynamics. Thus, we set the threshold to compute a predictability barrier as *PP*_lim_ =1 − (1 −0.25)^1/2^ =0.1340. Henceforth, we define predictability barrier as the number of weeks for which *PP* (*t*) =*PP*_lim_ and where the dynamics beyond this barrier can be considered unpredictable. Finally, note that the computation of *PP*(*t*)can also include parameter estimation uncertainty (20). Nonetheless, because we estimated the ARMA(2,2) parameters from the full time series (40 weeks) and we are dealing with a low-dimensional model, parameter uncertainty is expected to have a marginal effect (40); nonetheless, our estimates of *PP* (*t*)could be considered optimistic.

Finally, to test whether the severity of the initial outbreak (March–early May 2020) affected the ensuing cyclicity and predictability, we used previously estimated values of the basic reproduction number, *R*_0_, from death data at the county and state levels (30, 41); the time period for which these *R*_0_ values were estimated did not overlap with the time series used in the present study. The method for estimating these *R*_0_ values used the observed death counts and a statistical state-space modeling approach similar to our computation of *r*(*t*) in the present study. Also, the estimation of the *R*_0_ values in the previous studies was designed to factor out the effects of the timing of epidemic onset (higher spread rates occurred earlier in the epidemic) and population density (higher spread rates occurred at higher densities). Nonetheless, the estimates of the *R*_0_ values are directly comparable to *r*(*t*); they use the same type of data and methodology, but characterize different periods of the pandemic and different dynamical characteristics; see (30) for further technical details.

### Simulations

To help interpret the *r*(*t*) time series and investigate possible mechanisms underlying their cyclicity, we used a stochastic, discrete-time, age-structured Susceptible-Exposed-Infectious-Removed (SEIR) model, parameterized with published results (30). This simulation model tracks the epidemic on a daily time scale and explicitly includes the time period from infection to subsequent transmission (infectiousness), and from infection to death when the disease is reported. We modified the published model to explicitly separate a constitutive disease reproductive number, *R*_c_, from dynamic changes in the transmission rate that depend on the death count two weeks previously. In this way, we mimicked a susceptible population becoming aware of increases in the death toll and, following a 2-week delay for reporting and media attention, taking protective measures (individual behavioral responses and/or NPIs) (26, 27, 42). We set the transmission rate to

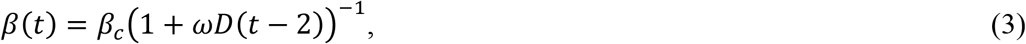

where *β*_*c*_ is the transmission rate corresponding to *R*_c_, *D* (*t* − 2)is the number of deaths two weeks previously, and *ω* scales how rapidly the transmission rate decreases with increases in *D* (*t* − 2). We selected this functional form to mimic the cyclicity in the observed data, although similar disease dynamics may be generated using other functions that decrease with *D* (*t* − 2). Our modeling approach is similar to that used by Weitz et al. (28), although our model explicitly incorporates the dependence of transmission and death on the number of days since infection, making it possible to compare our simulation results with real data. For further simulation details, see the *SI Appendix*.

The simulation model is built on the hypothesis that cyclicity is determined by differences in the constitutive and/or dynamic components of the transmission rate among jurisdictions. Our analyses, however, do not test this hypothesis directly. Instead, by comparing the simulated and real dynamics, we ask whether the hypothesis is plausible.

## Results

### Predictability and cyclicity at the county and state levels

Predictability measured by *PP*(*t*) varied substantially among counties and states (Fig. 2). For example, at the county level and for four-week-ahead forecasts, *PP*_4_ ranged from 0.03 to 0.72. This among-jurisdiction variation in *PP*(*t*) for any week *t* reflected high variation in the predictability barrier (Fig. 2a-b, *SI Appendix*, Fig. S4): counties had a median of 9 weeks (interquartile range 7-12 weeks), and states had a median of 8 weeks (5-11). *PP*_4_ – chosen to reflect the empirically found barrier of four weeks (see the *Introduction*) – characterizes the variation in predictability barrier among jurisdictions (*SI Appendix*, Fig. S5), and therefore we focus on *PP*_4_ throughout most of the remaining analyses.

**Fig. 2.**
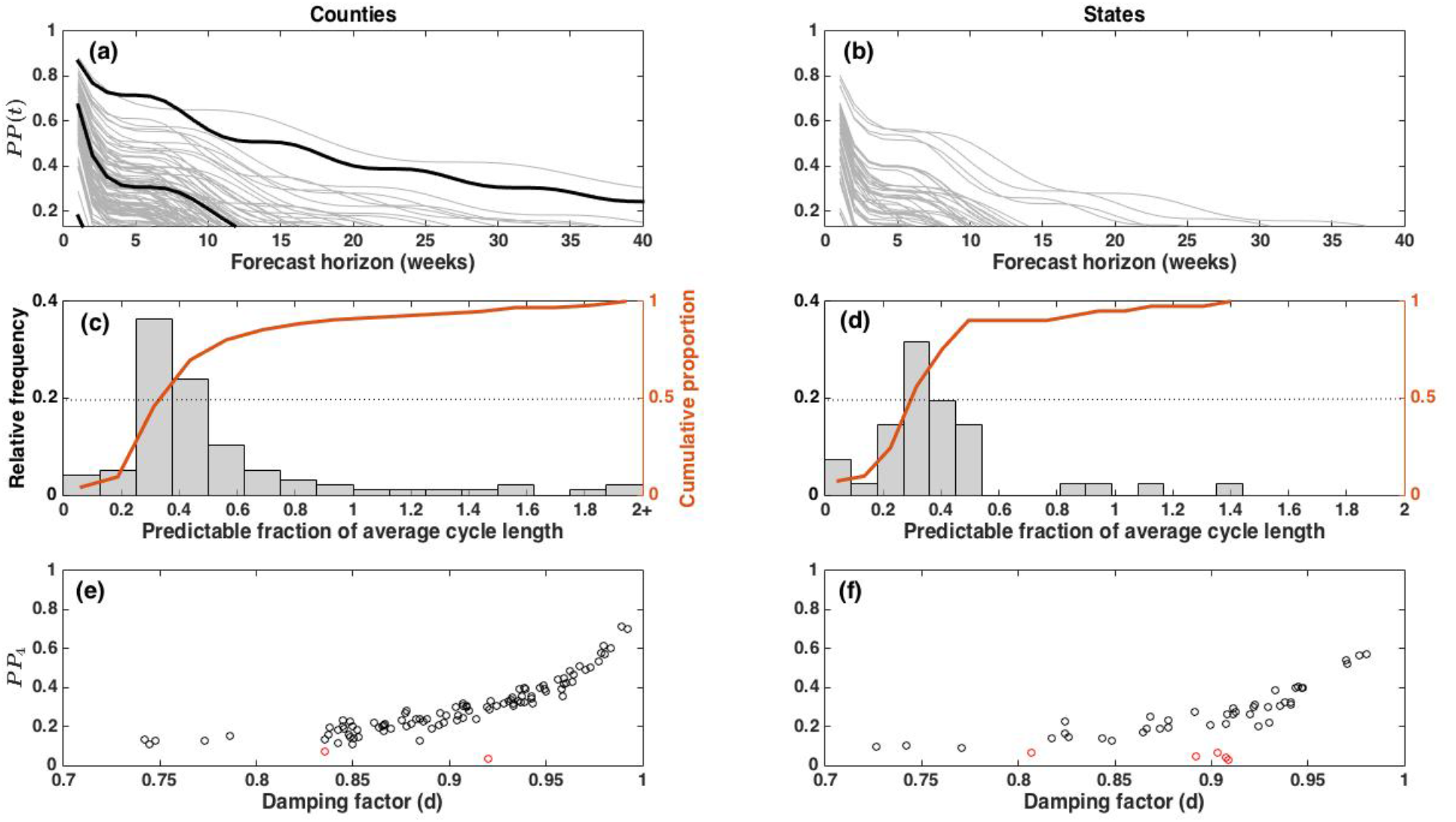
Predictive power and cyclicity. Predictive power, *PP*(*t*) (eq. 2), for **(a)** 98 counties and **(b)** 46 conterminous states with stationary dynamics for a forecast horizon ranging from 1 to 40 weeks. The lowest value of the y-axes in (a-b) is the threshold *PP*_lim_ = 0.134, used to compute the predictability barrier (cf. *SI Appendix*, Fig. S4). The three highlighted counties in (a) are those in Fig. 1 and Fig. 3: New York (five boroughs), New York; Maricopa County, Arizona; El Paso County, Texas. Panels (**c-d**) show the distribution of the estimated predictability barriers of jurisdictions with cyclic dynamics (cf. panels (a-b) and *SI Appendix*, Fig. S4) as a fraction of the average cycle length (counties: 23 weeks, states: 26 weeks), along with the cumulative proportion of jurisdictions. Finally, for both (**e**) counties and (**f**) states, *PP*_4_ was closely associated with the strength of cyclicity as measured by the damping factor *d*; the latter corresponds to the characteristic return time (38) for non-cyclic jurisdictions (red dots).

Of the 100 counties and 49 states, 96 and 41 showed cyclic dynamics in the stationary domain (*SI Appendix*, Fig. S6), and all analyzed jurisdictions had mean *r*(*t*) of nearly zero (not shown). The estimated period was similar at the county and state levels (*SI Appendix*, Fig. S7a,b): counties had a median of 23 weeks (interquartile range 20–29), and states had a median of 26 weeks (20–33). The damping factor (*d*) was also similar (*SI Appendix*, Fig. S7c,d): counties had median *d* = 0.91 (0.85–0.96), and states had median *d* = 0.91 (0.83–0.94).

Expressing the predictability barrier as a fraction of the median period (23 weeks and 26 weeks, see above) shows that for half the counties with stationary cyclic dynamics, at most 40% of a cycle is predictable, while at the state level it is 35% (Fig. 2c-d). Furthermore, only 10% of counties and 5% of states had a fully predictable cycle (‘wave’) or beyond. Second, we found a strong association between predictability and damping factor (Fig. 2e,f) (counties: Spearman’s *ϱ* = 0.83, *P* < 10^−10^; states: *ϱ* = 0.52, *P* = 0.0001); note that this result is not a mathematical inevitability: for example, near-random-walk dynamics are non-cyclic yet still imply high damping factors. In contrast to this association, we could not find a significant relationship between predictability and period (*SI Appendix*, Fig. S8), and therefore we will use the damping factor as a measure of cyclicity to investigate what causes the joint variation in cyclicity and predictability.

### Simulation results

The simulation model mimics the cyclic dynamics shown in the data (Fig. 3). Increases in cyclicity and predictability in the simulations are generated by increasing the constitutive reproduction number, *R*_c_. Because higher *R*_c_ values correspond to higher maximum values of *r*(*t*), more pronounced cyclicity and increased predictability occur when there is greater potential for rapid increases in disease spread rates. In the specific model realizations, increasing the *R*_c_ value from 1.4 (Fig. 3d) to 1.8 (Fig. 3f) increases *PP*_4_ from 0.11 to 0.55.

**Fig. 3.**
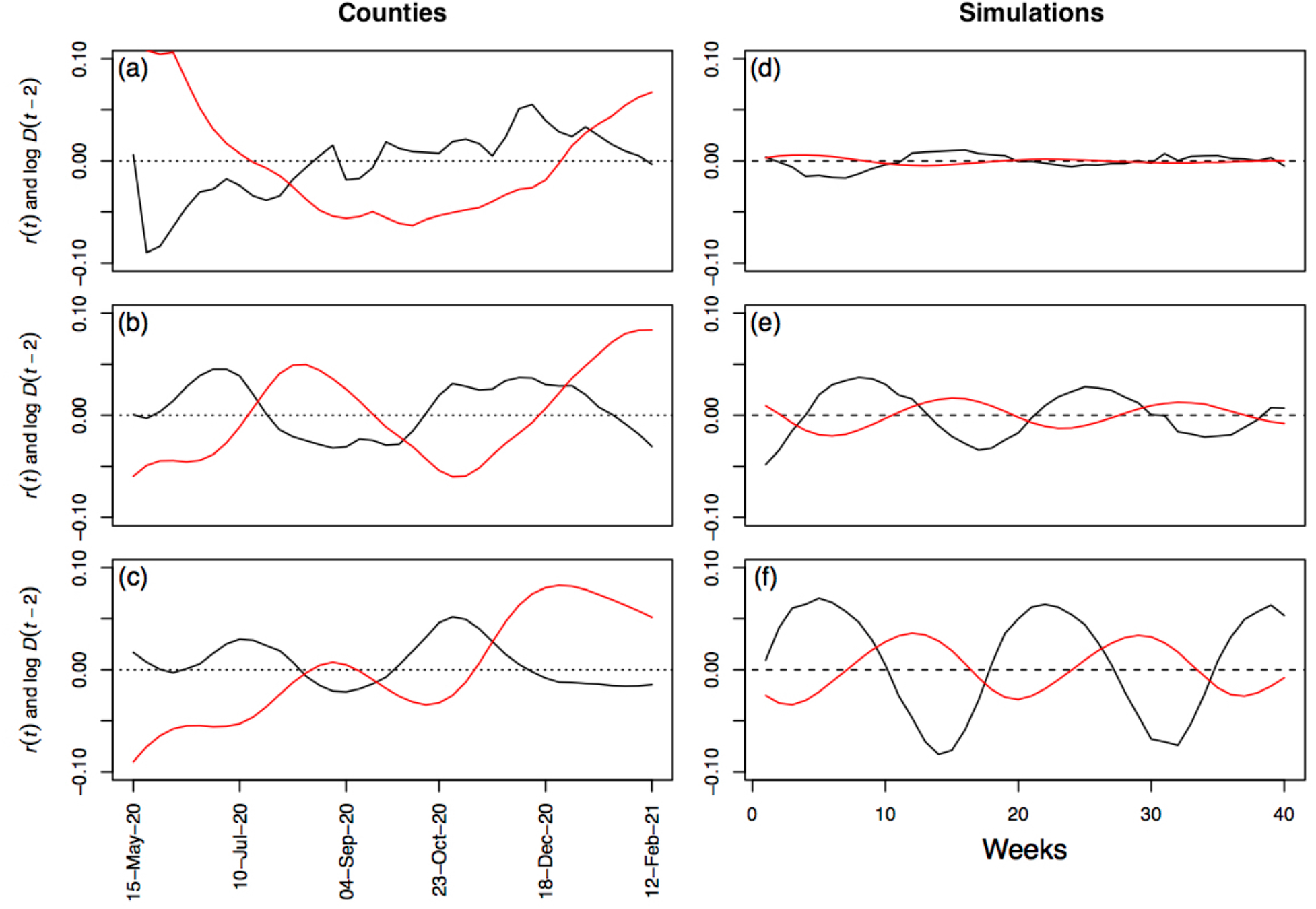
Comparison between *r*(*t*) estimated for three illustrative counties and for three simulated populations. The three counties are those in figure 1: (**a**) New York (five boroughs), New York, for which *PP*_4_ = 0.03, (**b**) Maricopa County, Arizona, with *PP*_4_ = 0.32, and (**c**) El Paso County, Texas, with *PP*_4_ = 0.72. For the simulations, *β*_*c*_ was selected to give three values of *R*_c_ (eq. 3): (**d**) *R*_c_ = 1.4, *PP*_4_ = 0.11, (**e**) *R*_c_ = 1.6, *PP*_4_ = 0.28, and (**f**) *R*_*c*_ = 1.8, *PP*_4_ = 0.55. In all panels, black lines give the estimates of *r*(*t*) and red lines give the z-transformed log number of deaths per week incorporating a 2-week time lag.

To compare with the county-level data, we simulated time series of 40 weeks using values of *R*_c_ randomly distributed between 1.4 and 1.8 (Fig. 4). Analyzing the simulated data in the same way as the real data, these simulations spanned the range of *PP*_4_ observed in the county data (Fig. 4a). In the simulations, the association between the damping factor *d* and *PP*_4_ (Fig. 4b) was very close to that found for the county data (Fig. 4e). The periods estimated from the simulated data were less variable than for the real data, although most fell between 20 and 30 weeks (Fig. 4c,f).

**Fig. 4.**
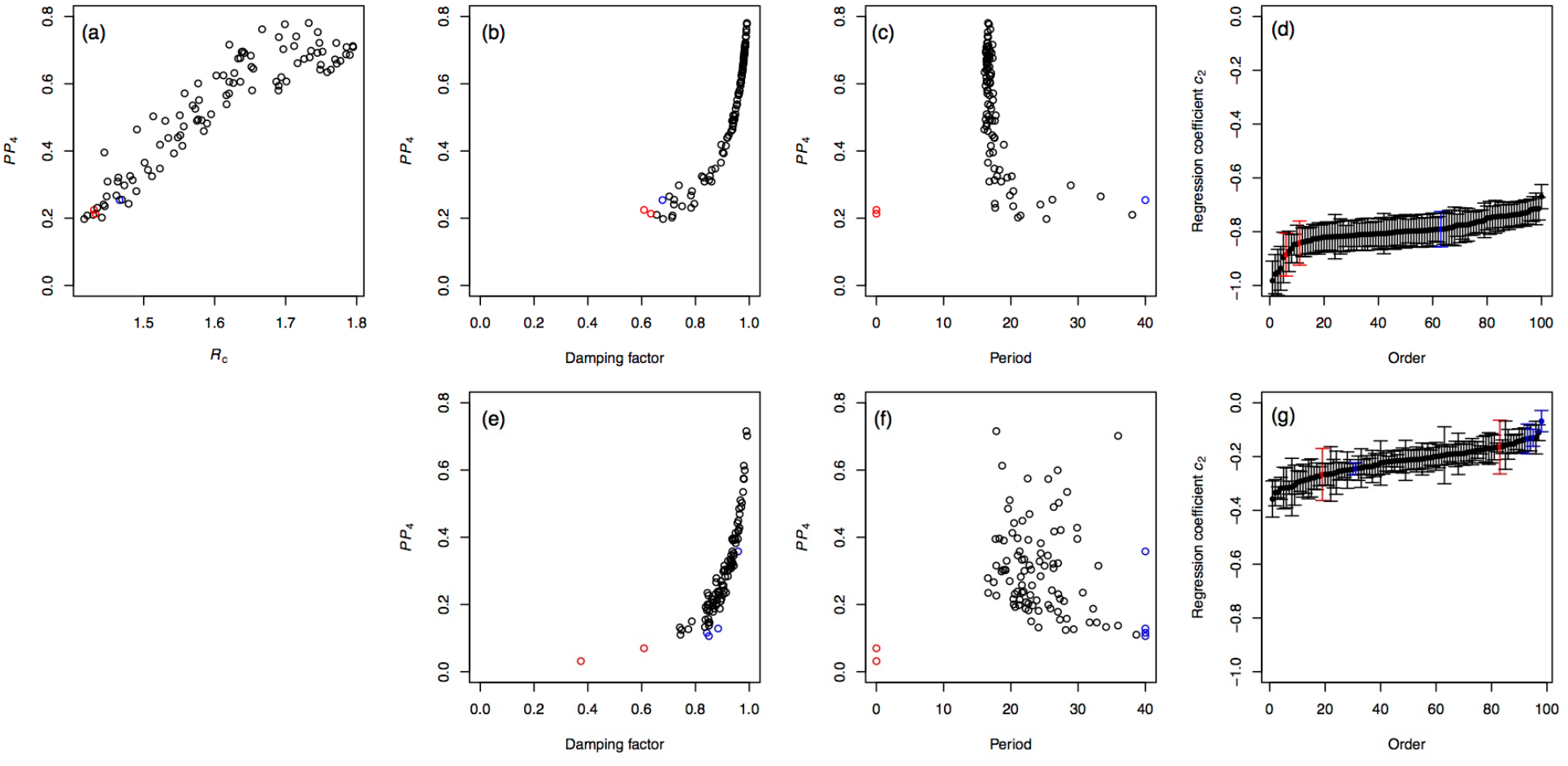
Comparison between dynamical characteristics of *r*(*t*) for simulated and real time series. One-hundred time series of *r*(*t*) were simulated for randomly selected values of the transmission rate *β*_*c*_ (eq. 3) to give values of *R*_c_ ranging from 1.4 to 1.8. (**a**) Predictability measured by *PP*_4_ increased with *R*_c_ implying that simulations in which the potential for increases in the spread rate were greatest (those with highest *R*_*c*_) had the most predictable dynamics. Simulated time series that were identified as non-cyclic are shown in red, and time series with period > 40 weeks are shown in blue, respectively. The association between predictability and cyclicity for (**b**) the simulated data was similar to that for (**e**) the real data. For simulated counties, (**c**) the period showed less variation than (**f**) the real data, although the overall relationship with *PP*_4_ was similar. No simulated time series was non-stationary, and the two non-stationary county time series are excluded because *PP* is undefined. Finally, (**d**) and (**g**) show the ranked regression coefficients for the effect of *D* (*t*−2) on *r t* for the simulated counties and county data, where bars give standard errors.

The key feature of the simulations generating cycles is the decrease in the transmission rate caused by increases in the death count two weeks beforehand (eq. 3). This feature of the simulation can be recovered statistically from the simulated time series by performing a conditional least-squares regression of *r*(*t*)against *r* (*t* − 1)and *D* (*t* − 2). For the 100 simulated counties, the regression coefficients ranged between −1 and −0.4 (Fig. 4d). For the county data, these regression coefficients ranged between −0.4 and −0.05 (Fig. 4g), and all but one (for a non-cyclic time series) are statistically significantly below zero (*p* < 0.05).

### R_0_ and variation in predictability

At both the county and state levels, the *R*_0_ values and *PP*_4_ were strongly negatively associated (Fig. 5a,b; counties: Spearman’s *ϱ* = -0.63, *P* < 10^−10^; states: *ϱ* = -0.52, *P* = 0.001): more severe initial outbreaks were followed by disease spread dynamics with lower predictability. This is the opposite pattern from what would be expected if high *R*_0_ values were followed by high constitutive *R*_c_ values; in the simulations, higher *R*_c_ values were associated with higher *PP*_4_ (Fig. 4a). These results imply that higher *R*_0_ values gave rise to ensuing dynamics with lower *R*_c_ values, suggesting that populations were constitutively more cautious in counties and states that had experienced a severe COVID-19 outbreak at the start of the pandemic.

**Fig. 5.**
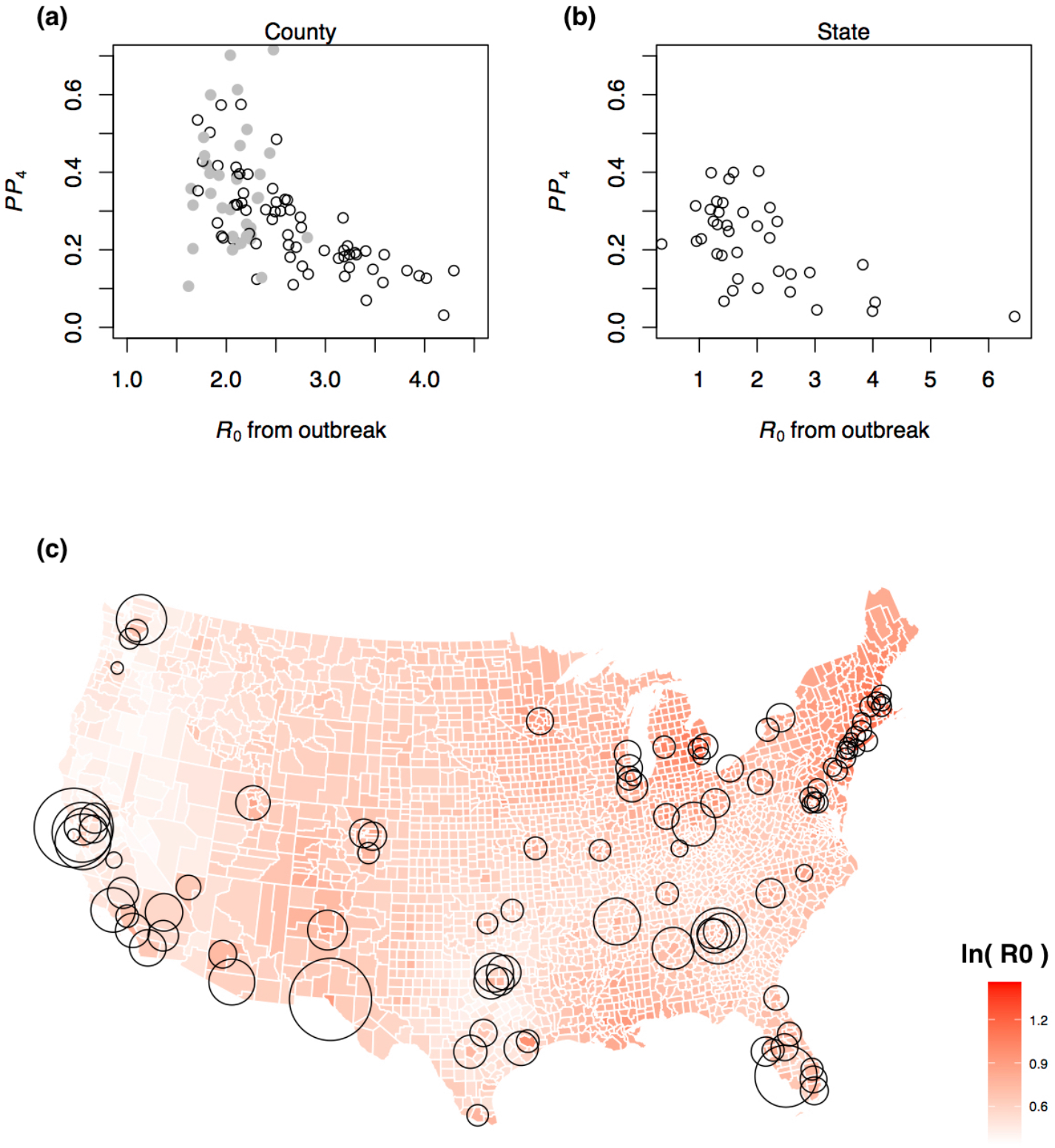
Effect of *R*_0_ on predictability. (**a–b**) Predictability as measured by *PP*_4_ is plotted against the *R*_0_ values estimated at the onset of the pandemic from death count records (30, 41). For county-level data, *R*_0_ values were computed for 124 counties, and those counties overlapping with the counties in the present study are shown as open points; the gray points correspond to counties for which values of *R*_0_ were interpolated using population density and geographical location. (**c**) The map gives the log-transformed county-level estimates of *R*_0_, originally ranging from 1.3 to 4.5 (white to dark red; (30)), with county-level estimates of *PP*_4_ depicted by circles, with circle diameter from smallest to largest corresponding to *PP*_4_ from 0.03 to 0.72.

Figure 5c overlays county estimates of *PP*_4_ on a map of the county estimates of *R*_0_ values from the initial outbreaks. A cluster of counties with low *PP*_4_ occurs along the northeastern coast where *R*_0_ values were high, while counties with high *PP*_4_ and pronounced cyclicity occur in southern states and in California.

## Discussion

The COVID-19 pandemic has stimulated the development of numerous quantitative models to help understand and forecast disease dynamics, and to assist public health decision-making (e.g. 12, 22, 43). Rather than develop methods for making predictions, in this study we have focused on the inherent unpredictability of COVID-19 dynamics. Our goals have been both to address the limits to which predictions are possible for communicable diseases like COVID-19, and to understand the dynamical characteristics of epidemics that make predictions more or less accurate.

We found considerable variation in predictability among jurisdictions (Fig. 2, *SI Appendix*, Fig. S4), as also found by (15). In contrast to (15), however, we found that for the majority of analyzed counties and states, the predictable fraction of a cycle (that is, an outbreak in (15)) is much less than one (Fig. 2). Our estimated cycle lengths are in good agreement with previous findings (10, 11). In addition, we show that predictability is strongly related to the rate at which cycles are damped, with weakly damped cycles giving regular patterns in the data that allow predictions: this rate of cycle damping has been largely neglected in previous analyses. Finally, we show that protective measures against COVID-19 reduced both the cyclicity and predictability of disease dynamics. Thus, variation in cyclicity and predictability among jurisdictions gives valuable information about factors governing the dynamics of COVID-19.

In analyses of forecast accuracy, single studies and reviews of the many studies forecasting COVID-19 dynamics have focused on identifying the best forecasting methods (e.g. 12, 19). Our analyses of inherent unpredictability focus on how much information is available in a time series, rather than the ability of a model to fit the time series and make forecasts. Therefore, our estimates of the limits to forecasts in principle should apply to all forecasting models. Furthermore, our demonstration of the high variation in predictability among time series from different counties and states in the USA implies that the ability to forecast COVID-19 likely depends more on the dynamics in a particular dataset than on the forecasting methods used.

Our simulation model showed that cyclic dynamics similar to those observed in the county and state data can be mimicked when changes in the transmission rate occur as a 2-week delayed response to increases in the death toll. We acknowledge that this is not categorical evidence that time-delayed changes in the transmission rate in response to death counts are responsible for the cycles, because any form of cyclicity in *D*(*t*) will drive cyclicity in *r*(*t*).Nonetheless, this pattern is consistent with the hypothesis under which the simulation model was built. The simulation model shows the plausibility of the hypothesis that more pronounced cyclicity occurs in jurisdictions with higher constitutive *R*_c_ values, because a higher *R*_*c*_ allows more rapid changes in the transmission rate that are necessary to generate cycles. Finally, jurisdictions that experienced severe outbreaks at the onset of the pandemic, measured by high values of *R*_0_ before widespread public protective measures were put in place, had less cyclic and less predictable COVID-19 dynamics in the subsequent period before vaccination became widespread. The association between a high *R*_0_ value and lack of predictability suggests that a severe initial outbreak led to high levels of constitutive protective measures which individuals took to reduce disease transmission. Moreover, the variation in predictability had a clear geographical pattern, with many counties having unpredictable dynamics occurring in the Northeast (Fig. 5).

The hypothesis embodied by our simulation model is that cyclicity arises from protective measures people take in response to rising death tolls (cf. 27), that is, a negative feedback loop much like “predator-prey” dynamics in ecology which has recently attracted increased attention in epidemiology (26, and references therein). Because death tolls are highly correlated with case counts, human responses could equally depend on the awareness of rising cases, reports in the media, word-of-mouth, etc. Maps of current cases and deaths from COVID-19 were publicly available throughout the time period we analyzed, and reports of case counts occurred regularly in the news. Some responses to increased spread of COVID-19 were taken by policy-makers, such as mask mandates and restaurant closures. Other responses were taken by individuals to reduce contact and abide mandates. We have shown that if the ‘background’ constitutive transmission rate of COVID-19 is high, then the human response to increasing disease spread will generate pronounced cyclic dynamics. In contrast, if the constitutive transmission rate is kept low, then cycles do not appear, because the disease dynamics are not as responsive to changes in protective measures. This implies that lack of cyclicity and predictability are caused by people continuously take greater precautions against COVID-19, rather than showing an on-and-off response to changes in death tolls or case counts.

Our analyses are based on observed disease dynamics to make inferences about differences in protective measures taken by citizens of states and counties. There has been considerable research effort to assess attitudes, such as surveys on mask use (44) and vaccination hesitancy (45), and to identify effective proxies of protective behaviors, such as analyses of government policies (31) and changes in individual movement patterns using cell-phone signals (46). While acknowledging the value of these studies, our approach of analyzing the dynamics of COVID-19 focuses on the effects of protective behaviors, rather than the protective behaviors themselves. Even though our approach cannot make a mechanistic link between behaviors and dynamics, it nonetheless gives insight into differences in how COVID-19 was experienced by different jurisdictions.

Our explanation for the joint variation in cyclicity and predictability is a hypothesis that is consistent with our statistical evidence. Direct evidence is a challenge, however, because variation among jurisdictions in the constitutive protective measures that individuals take are hard to document. Nonetheless, the remarkable negative association between predictability and *R*_0_ (Fig. 5) suggests differences in personal protective measures among jurisdictions. Before performing our analyses, we hypothesized that *R*_0_ values would be positively associated with predictability, because a high *R*_0_ value implies the potential for rapid increases in disease spread if protective measures were dropped. Our finding of a negative association suggests that populations experiencing severe initial outbreaks saw a fundamental shift in later transmission rates. An alternative explanation for this shift is that the initial outbreak generated sufficient acquired immunity to reduce future transmission rates (11). Arguing against this explanation, however, is that during the period we analyzed the number of COVID-19 cases as a proportion of the population ranged from 1–14% among counties and 2–13% among states. Furthermore, there was no relationship between the cumulative per capita number of cases and *PP*_4_ for either county (Spearman’s *ϱ* = 0.12. *p* = 0.22) or state data (*ϱ* = 0.23. *p* = 0.11). Even though cases were likely under-reported, serological studies show that, for example, the proportion of the adult population in New York City having contracted COVID-19 between 19 April and 5 July, 2020, was approximately 20% – similar results have been found for metropolitan France (approximately 15% of adults by January 2021) – which is likely not high enough to affect the subsequent predictability of the dynamics (47, 48). It is also possible that cyclicity was driven by successive SARS-CoV-2 variants each with higher transmission rates (25). While different variants are associated with differences in *R*_0_ among jurisdictions at the start of the pandemic (30), and successive variants were more transmissible (49), the successive variants spread geographically quickly throughout the conterminous USA. While new variants might have added to the broad pattern of cyclicity of COVID-19 in the USA, we cannot think of how new variants could explain the negative association between *R*_0_ values and subsequent cyclicity. Given that acquired immunity and SARS-CoV-2 variants are unlikely explanations for the negative association between *R*_0_ and predictability across jurisdictions, our best alternative is changes in protective measures taken by individuals.

What are the implications of our findings for decision-making in public health emergencies? The USA experienced repeated waves of COVID-19 after the initial spread of the pandemic, and these waves caused large numbers of infections and deaths. Nonetheless, after the initial rapid outbreaks, the spread rates were lower (compare the results in (30) to *SI Appendix*, Fig. S1). This suggests that steps taken by policymakers and individuals to reduce transmission rates – such as mask wearing, social distancing, and other NPIs – were effective. Indeed, the lack of predictability can be viewed as a consequence of the successful maintenance of low transmission rates. If COVID-19 spread rates are predictable, it means that protective measures have been dropped and therefore have to be restarted. Although the consequence of a population taking continuous protective measures is lack of predictability, lack of predictability itself is an indicator of effective transmission management. Our results further indicate that one of the first epidemic-related metrics computed at the early stages of an epidemic, namely *R*_0_, allows anticipating the predictability of the ensuing dynamics (Fig. 5). For outbreaks of newly emerged diseases this information could be complemented by jurisdiction-specific data indicating how well NPIs in the past have been successful, in terms of swift implementation and adherence by the population (e.g. 31, 43): this would give information about how strongly protective measures will affect disease dynamics and consequently their predictability. Finally, all our results are similar at the county and state levels, implying that at the onset of outbreaks, information from different jurisdictional levels could be helpful to gauge the limit to forecasting accuracy.

The human response to disease spread likely affects its predictability, and the pandemic might be similar to stock markets in which unpredictability is generated by human behavior (50). We should anticipate that future pandemics will be similarly unpredictable if they elicit widespread behaviors to reduce transmission. Unpredictability is just a by-product of positive steps that people take to protect themselves and others.

## Data Availability

All data used in the present work have been previously published.

## Data availability

After acceptance of the manuscript, data that support the findings of this study will be uploaded on figshare.

## Code availability

After acceptance of the manuscript, R code for the analyses will be uploaded on figshare.

## Acknowledgments

This work was supported by NASA-AIST-80NSSC20K0282 (A.R.I).

## Author contributions

C.B. and A.R.I. designed the study and methodology; C.B. and A.R.I. performed the analyses; C.B. and A.R.I. wrote the manuscript.

## Competing interests

The authors declare no competing interests.

## Notes

### Competing Interest Statement

The authors have declared no competing interest.

### Funding Statement

This study was funded by: NASA-AIST- 80NSSC20K0282

