## SupplementaryInformation for "Differences in COVID-19 cyclicity and predictability among U.S. counties and states reflect the effectiveness of protective measures"

**The *SI Appendix* contains:**

- **Supplementary Figures S1–S8**
- **Additional methods information**

#### Supplementary Figures

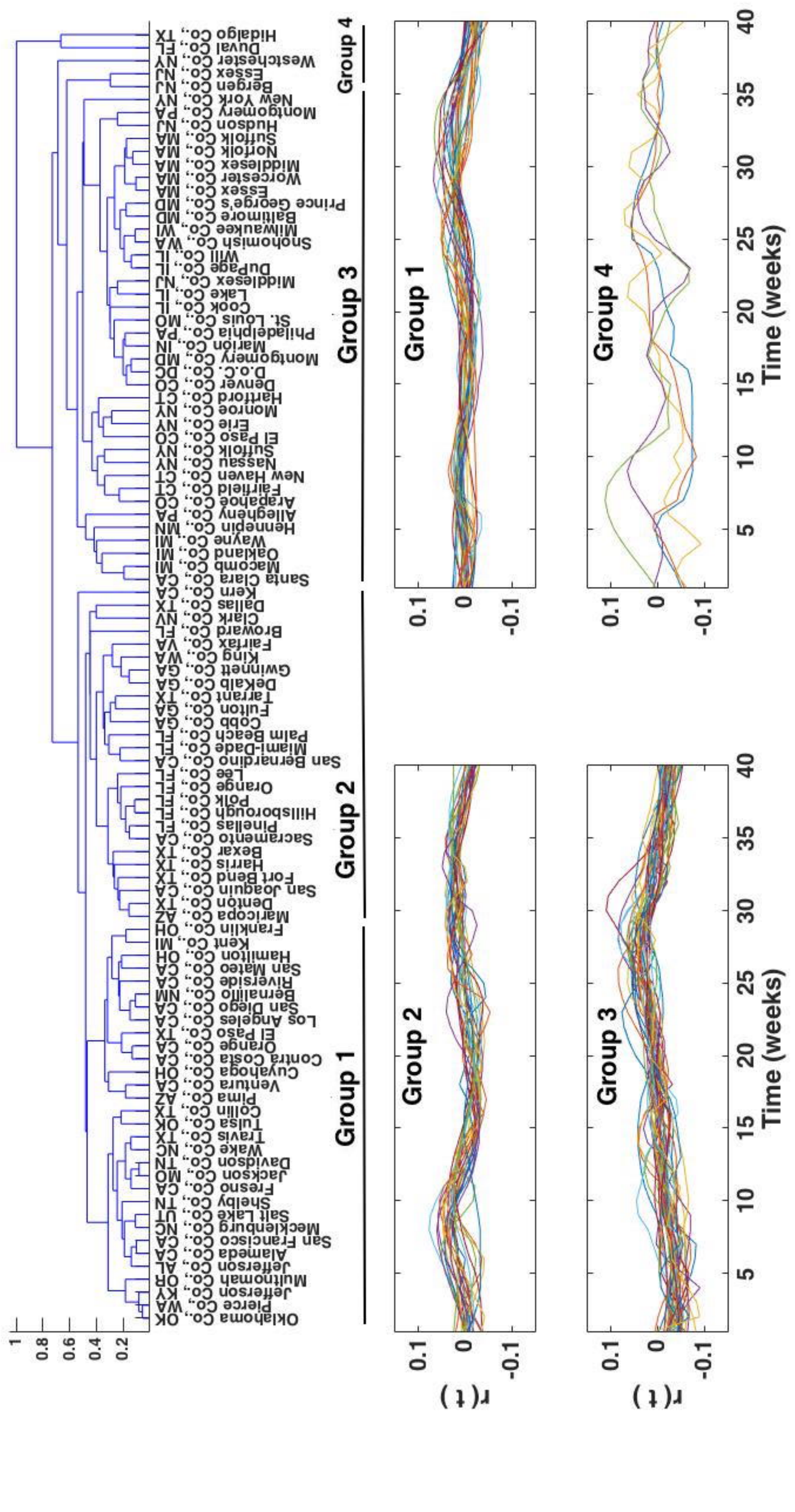


**Fig. S1. Estimated *r(t)* time series at the county level**. The estimated *r*(*t*) time series of the 100 analyzed counties are grouped according to the similarity of the dynamics. The similarity has been assessed using a dynamic time warping algorithm (1, 2), and for illustrative purposes four groups have been used.

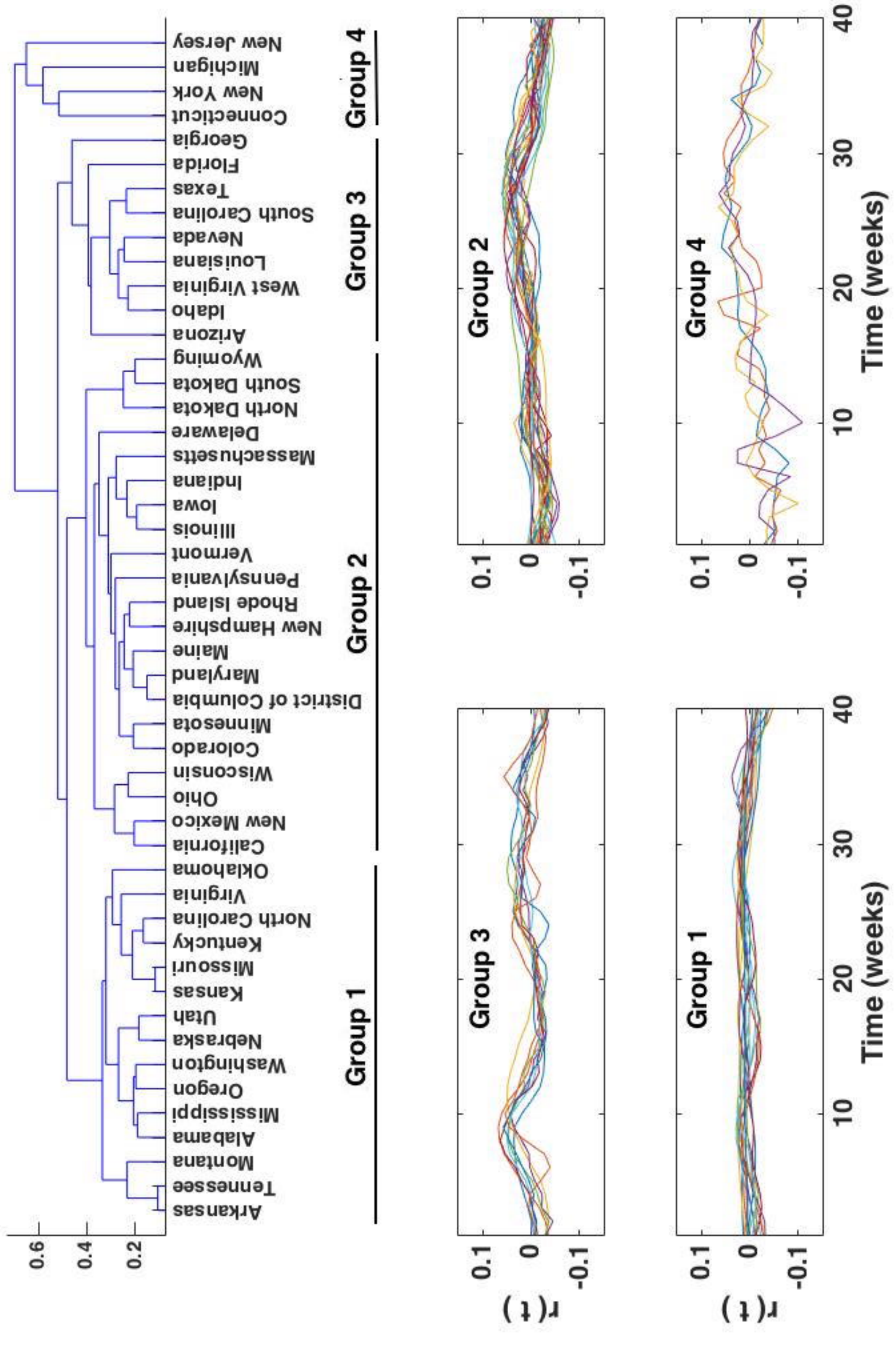


**Fig. S2. Estimated *r(t)* time series at the state level**. The estimated *r(t)* time series of the 49 analyzed states are grouped according to the similarity of the dynamics. The similarity has been assessed using a dynamic time warping algorithm (1, 2), and for illustrative purposes four groups have been used.

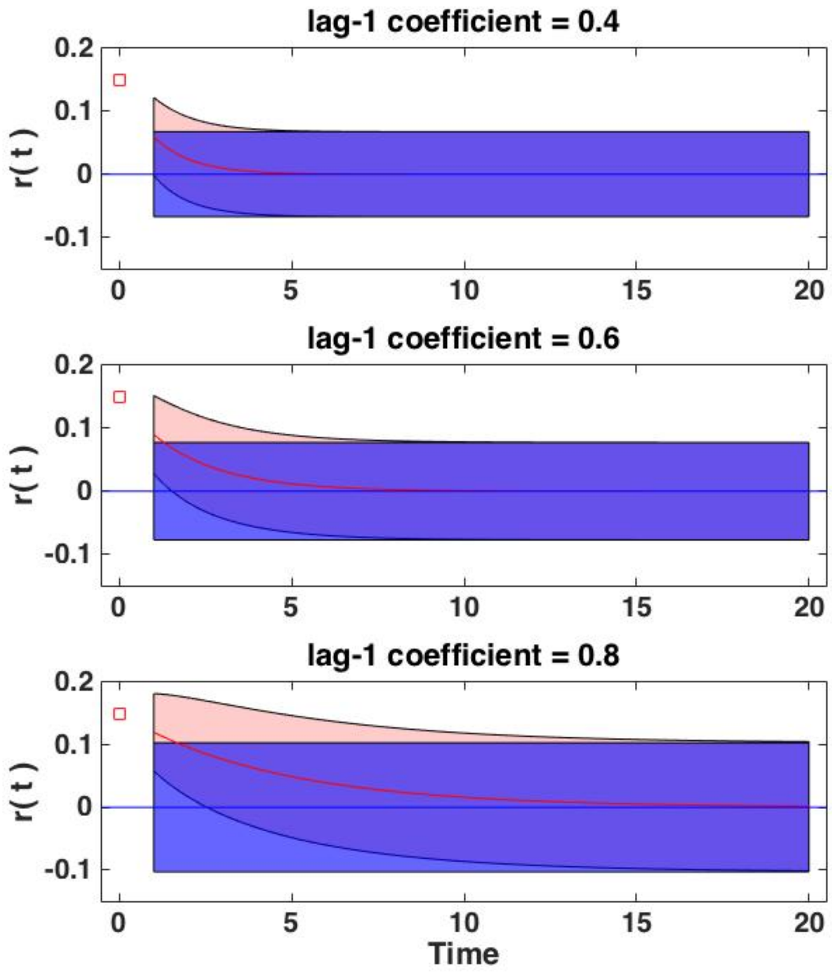


**Fig. S3. Illustrative visualization of the transition distribution overlapping with the stationary distribution.** For three illustrative AR(1) processes, the time-dependent transition distribution and the stationary distribution are shown, which are used to compute the predictability measure predictive power ($PP(t)$, eq. 2); the variances here are shown as 95% confidence intervals, and the red square is an arbitrary initial state. The quicker the transition distribution is 'absorbed' by the stationary one, the faster predictability is said to be lost. Simulation parameters: lag-1 coefficients are given in the panel titles; $r(t = 0)$ = 0.15 and var($r$) = 0.001 for all three AR(1) processes.

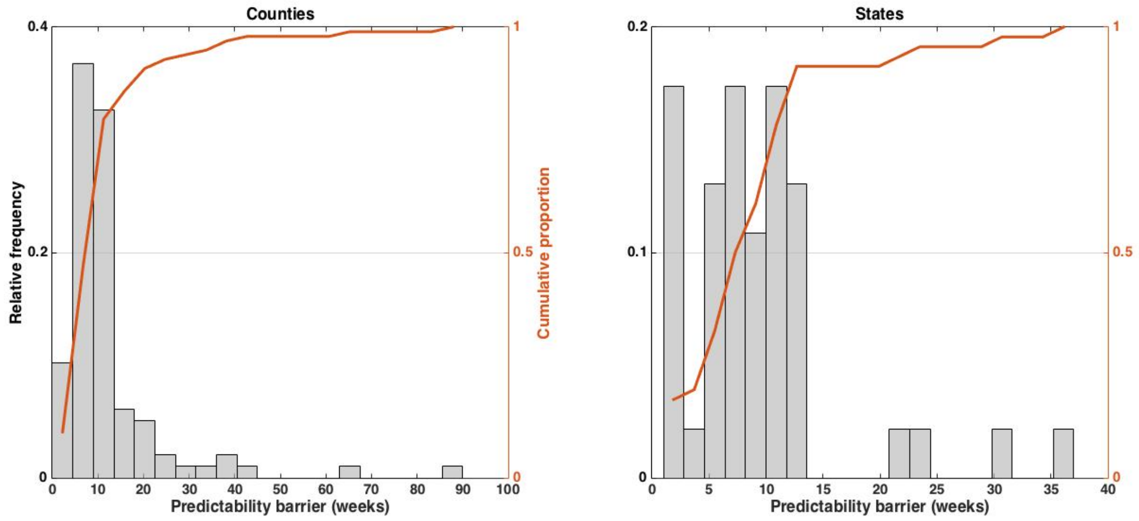


**Fig. S4. Estimated predictability barrier**. Shown are the distributions of the estimated predictability barriers (in weeks) at the county and state levels, along with the cumulative proportion of the respective jurisdictions.

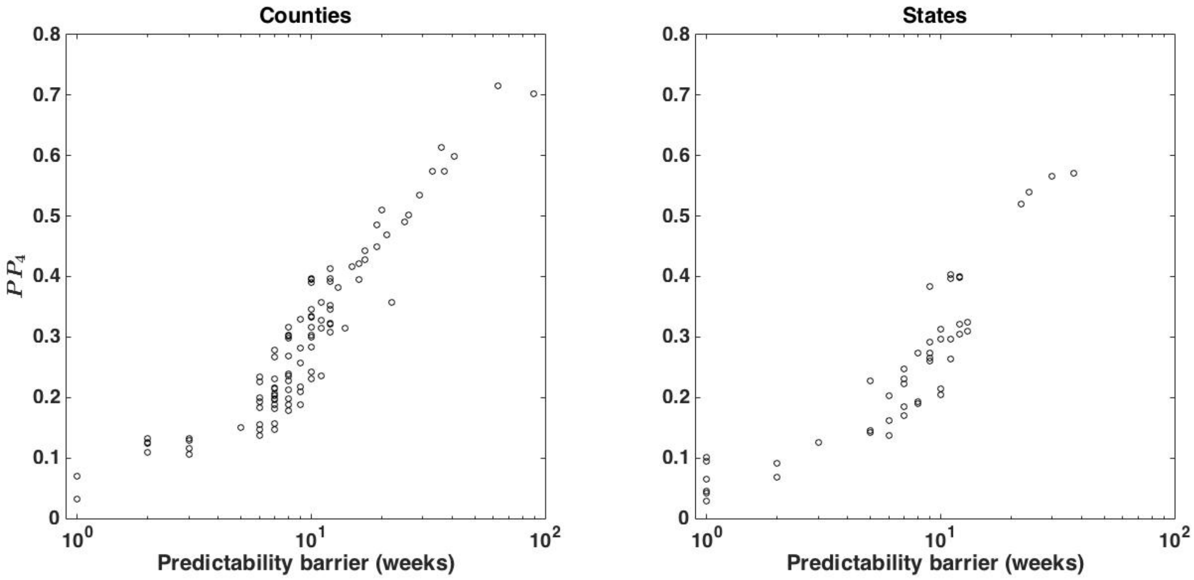


**Fig. S5. *$PP_4$* as a proxy for the predictability barrier**. The panels show that predictive power at time $t$ = 4 ($PP_4$) is a good proxy for the predictability barrier (in weeks), at both the county and state levels.

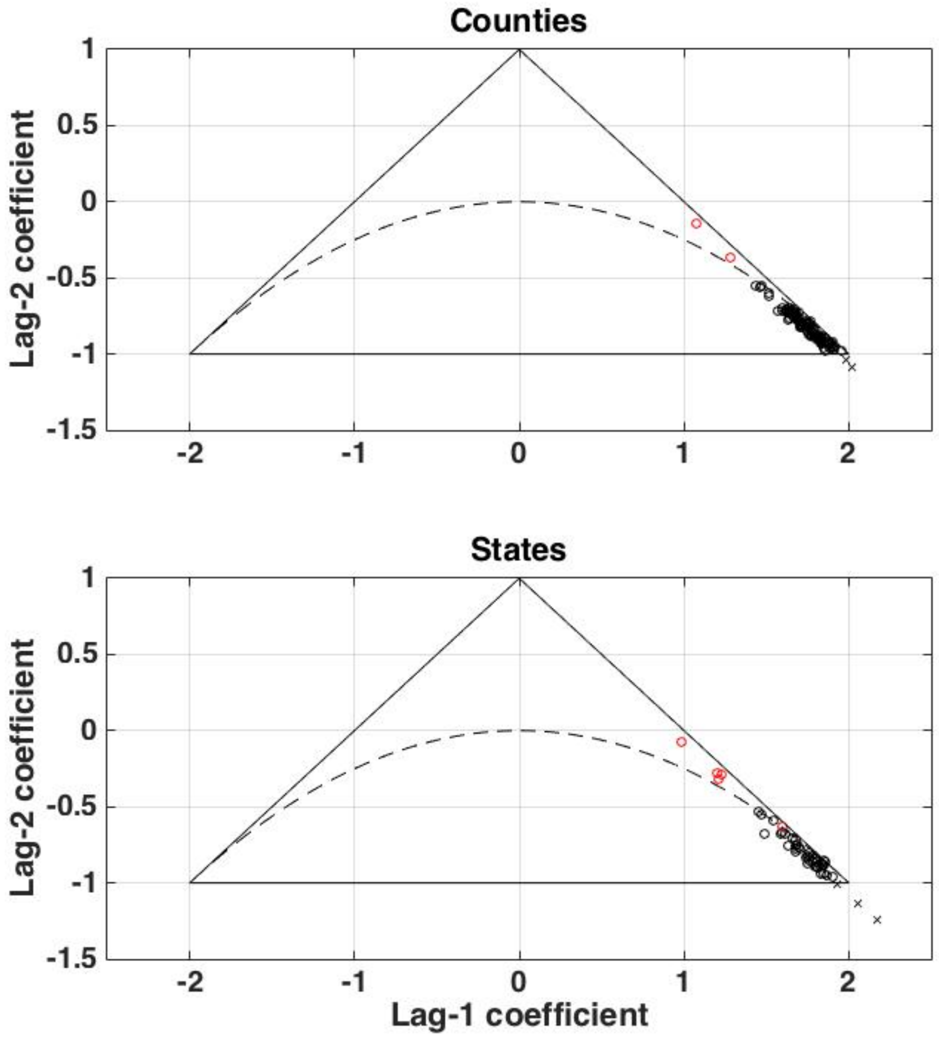


**Fig. S6. Estimated autoregressive model parameters**. Both panels show the estimated parameter pairs, i.e. the autoregressive lag-1 and lag-2 coefficients of the ARMA(2,2) model (eq. 1) fit to the $r(t)$ time series at the county level (cf. Fig. S1) and state level (cf. Fig. S2). Parameter combinations outside the triangle (marked with x's) imply a non-stationary process. Parameter combinations inside the triangle imply a stationary process, of which combinations below the parabola imply oscillatory dynamics (marked in black); estimated pairs implying non-oscillatory stationary dynamics are marked in red. The estimated lag-2 coefficients are approximately a linear function of the lag-1 coefficients: for counties, $b_2 \approx 0.80 - 0.93b_1$; for states, $b_2 \approx 0.87 - 0.97b_1$.

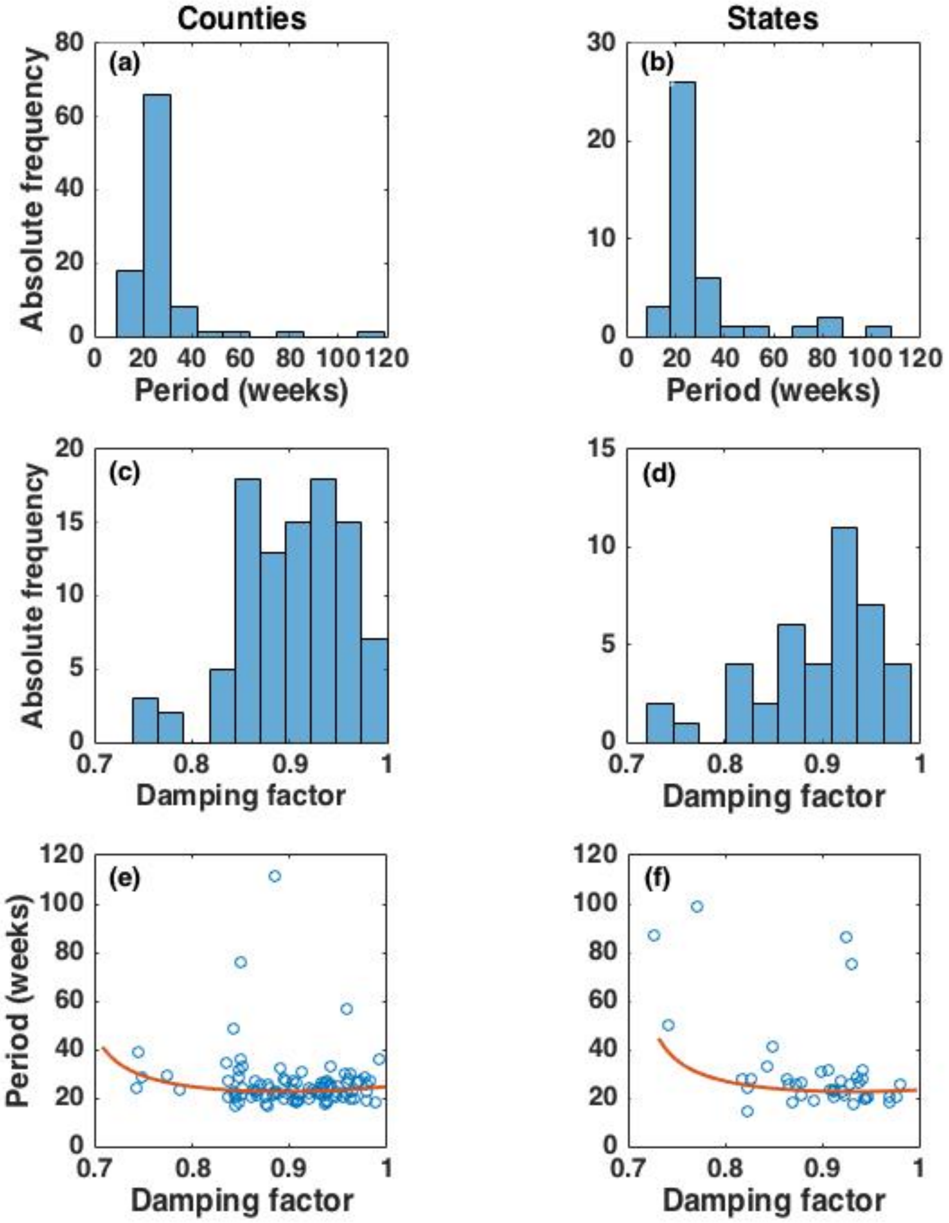


**Fig. S7. Estimated average cycle length and damping factor**. The two columns show, at the county and state level, **(a-b)** the distribution of the estimated average cycle length (period), **(c-d)** the distribution of the estimated damping factor, and **(e-f)** a scatter plot of the two measures. In the scatter plots, the red line was computed using the regression line for the two highly correlated AR(2) parameters $b_1$ and $b_2$ (cf. Fig. S4).

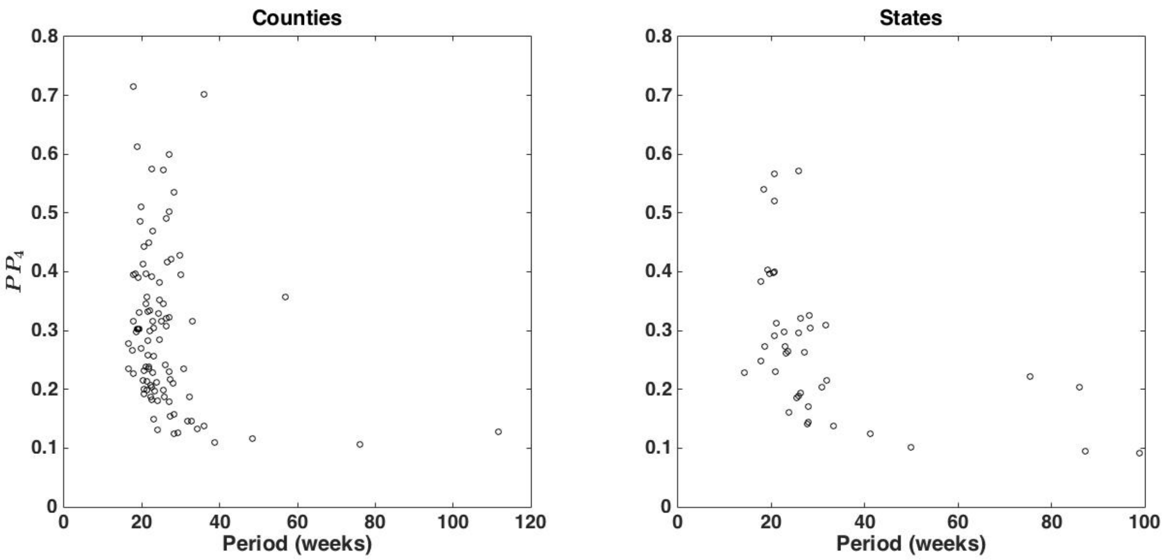


**Fig. S8. Predictability versus period**. The panels show, at the county and state levels, the estimated predictability as measured by predictive power at time $t = 4$ ($PP_4$) and the average cycle length (period), at both levels lacking a significant association.

### Additional methods information

*Predictive power for an* ARMA(2,2) *process*

The variance of the transition distribution and the variance of the stationary distribution of the ARMA(2,2) model (eq. 1 in the main text) must be calculated to compute the predictability measure predictive power ($PP(t)$, eq. 2). This can be done by recasting the ARMA(2,2) model as a vector autoregressive process of order one, that is, a 4-dimensional VAR(1); (3). To this end, the ARMA(2,2) model parameters populate the two jurisdiction-specific matrices

$$\mathbf{C} = \begin{bmatrix} b_1 & b_2 & a_0 & a_1 \\ 1 & 0 & 0 & 0 \\ 0 & 0 & 0 & 0 \\ 0 & 0 & 1 & 0 \end{bmatrix},$$
$$\mathbf{\Sigma} = \begin{bmatrix} \sigma^2 & 0 & \sigma^2 & 0 \\ 0 & 0 & 0 & 0 \\ \sigma^2 & 0 & \sigma^2 & 0 \\ 0 & 0 & 0 & 0 \end{bmatrix},$$

where $\mathbf{\Sigma}$ is a variance-covariance matrix, and $\sigma^2$ is the variance of the original univariate ARMA(2,2) process (eq. 1). The covariance matrix of the stationary distribution ($\mathbf{V}_\infty$) in vectorized form is defined as

$$\mathrm{vec}\left(\mathbf{V}_{\infty(\mathrm{VAR})}\right) \coloneqq (\mathbf{I} - \mathbf{C} \otimes \mathbf{C})^{-1} \mathrm{vec}(\mathbf{\Sigma}),$$

where vec(·) is the vec-operator, $\mathbf{I}$ is an identity matrix of appropriate size, ⊗ denotes the Kronecker product, and subscript VAR refers to the recast model. To compute the variance of the stationary distribution (as well as the transition distribution; see below) of the original univariate ARMA(2,2) process ($\mathbf{V}_{\infty(\mathrm{ARMA})}$, which is a scalar), the two additional vectors

$$\mathbf{j} = [1 \quad 0 \quad 0 \quad 0],$$
$$\mathbf{h} = [1 \quad 0 \quad 1 \quad 0]$$

are needed, so that (superscript T means the transpose)

$$\mathbf{V}_{\infty(\mathrm{ARMA})} = \mathbf{j}\, \mathbf{V}_{\infty(\mathrm{VAR})}\, \mathbf{j}^{\mathrm{T}}.$$

The time-dependent variance of the transition distribution of the original univariate ARMA(2,2) process ($\mathbf{V}_{\mathrm{t(ARMA)}}$, which is a scalar) can be computed as

$$\mathbf{V}_{\mathrm{t(ARMA)}} = \sum_{i=0}^{t-1} \boldsymbol{\Phi}_{\boldsymbol{i}}\, \sigma^2\, \boldsymbol{\Phi}_{\boldsymbol{i}}^{\mathrm{T}},$$

where the matrix $\boldsymbol{\Phi}_{\boldsymbol{i}} \coloneqq \mathbf{j}\, \mathbf{C}^{i}\, \mathbf{h}^{\mathrm{T}}$; further technical details can be found in (3). The transition distribution gives the evolution of the process through time, starting at a specific point $r_0$ at time $t_0$ (both potentially multi-dimensional) when the variance $\mathrm{var}(r_0) = 0$. We calculated the variance of the transition distribution for eq. 2 without considering spatial correlation, $\mathrm{cor}(\delta_i(t), \delta_j(t))$. The calculations are still correct for the marginal transition distribution, that is, the transition distribution for a single jurisdiction that does not depend on the other ones. Further, the differences in the mean spread rate among jurisdictions, $b_{0,i}$ (eq. 1), can also be ignored, because all estimates were nearly zero (*Results* in the main text).

*Simulation model*

The simulation model was designed to determine the plausibility of the hypothesis that changes in the transmission rate of COVID-19 in response to death counts could explain patterns of cyclicity and predictability observed in state and county data. The simulation model is a modification of that presented in (4). The simulation model tracks the epidemic on a daily time scale and explicitly includes the time period from infection to subsequent transmission (infectiousness), and from infection to death; therefore, it is akin to a SEIR model.

The simulation model tracks the number of infected individuals on day *t* who were infected $\tau$ days previously, $I(t;\tau)$. After 25 days, they are all assumed to be recovered or dead. The relative daily infectiousness of an infected individual, $p(t)$, is given by a Weibull distribution with mean 7.5 days and standard deviation 3.4[6] (Supplementary Fig. 3a in (4)). For an individual who dies, the day of death, $d(t)$, is given by a Weibull distribution with mean 18.5 days and standard deviation 3.4[6] (Supplementary Fig. 3b in (4)). Finally, the time between initial infection and diagnosis, $h(t)$, is assumed to be log-normally distributed with mean 5.5 days and standard deviation 2.2[8] (Supplementary Fig. 3c in (4)).

Deaths occur according the infection age category of individuals, $I(t;\tau)$, so that the probability of death of individuals that had been infected $\tau$ days earlier is $(1 - s)d(\tau)$. Here, *s* is the overall survival probability, assumed to be $s$ = 98%; changes in this assumption had little effect on the simulation results. Once an individual dies, they are removed from the pool of individuals. These individuals are summed to calculate the daily death count, $D(t)$.

On day $t$, the number of new infections is $\beta(t)S(t)\exp\big(\alpha(t)\big)\sum_\tau p(\tau)I(t;\tau)$, where $\beta(t)$ is the transmission rate and $p(\tau)$ scales the transmission rate by the infectiousness of individuals infected $\tau$ days earlier, $I(t;\tau)$. Here, we let $\beta(t) = \beta_c\big(1 + \omega D(t-14)\big)^{-1}$, so that an increasing number of deaths two weeks beforehand decreases the transmission rate below the constitutive transmission rate of $\beta_c$; this transmission rate function is the same as in eq. 3 in the main text, albeit here expressed on a daily timescale. The lognormal random variable $\exp\big(\alpha(t)\big)$ is included to represent environmental variation; its variance was selected to mimic the variation in $r(t)$ observed in the real data. Note that the simulations produce values of $r(t)$ that we analyzed directly, rather than estimating $r(t)$ by applying a Kalman smoother to the simulated values of $D(t)$.

Although the time series were generated on a daily time scale, simulated data were subsequently aggregated to weeks for analyses. The analyses of $r(t)$ were performed on time series of length 40 weeks. The results of the analyses depended on the initial number of infected individuals and the week at which the analyses were initiated. For the analyses in the main text, we initially simulated 40 weeks of data starting with $I(t;\tau) = 10^{-6}$. Other parameters were $\omega = 10^5$ and the variance of $\alpha(t) = 0.03$.

We analyzed each simulated time series separately, unlike the real time series which were all analyzed simultaneously. Furthermore, fits using an ARMA(2,2) were unstable for time series of length 42, and therefore we fit the simulated time series using an AR(2) model, estimating $PP_4$, the damping factor $d$, and average cycle length (period) from the autoregressive coefficients in the same way as the real data.

#### References *SI Appendix*


1. M. Müller, "Dynamic Time Warping" in *Information Retrieval for Music and Motion*, (Springer, 2007), pp. 69–84.
2. Q. Wang, *Dynamic Time Warping (DTW)* (2021) (September 8, 2021).
3. H. Lütkepohl, *New Introduction to Multiple Time Series Analysis* (Springer, 2005).
4. A. R. Ives, C. Bozzuto, Estimating and explaining the spread of COVID-19 at the county level in the USA. *Commun Biol* **4**, 1–9 (2021).
